# Multimodal Data Hybrid Fusion and Natural Language Processing for Clinical Prediction Models

**DOI:** 10.1101/2023.08.24.23294597

**Authors:** Jiancheng Ye, Jiarui Hai, Jiacheng Song, Zidan Wang

## Abstract

**Objective:** To propose a novel approach for enhancing clinical prediction models by combining structured and unstructured data with multimodal data fusion.

**Methods:** We presented a comprehensive framework that integrated multimodal data sources, including textual clinical notes, structured electronic health records (EHRs), and relevant clinical data from National Electronic Injury Surveillance System (NEISS) datasets. We proposed a novel hybrid fusion method, which incorporated state-of-the-art pre-trained language model, to integrate unstructured clinical text with structured EHR data and other multimodal sources, thereby capturing a more comprehensive representation of patient information.

**Results:** The experimental results demonstrated that the hybrid fusion approach significantly improved the performance of clinical prediction models compared to traditional fusion frameworks and unimodal models that rely solely on structured data or text information alone. The proposed hybrid fusion system with RoBERTa language encoder achieved the best prediction of the Top 1 injury with an accuracy of 75.00% and Top 3 injuries with an accuracy of 93.54%.

**Conclusion:** Our study highlights the potential of integrating natural language processing (NLP) techniques with multimodal data fusion for enhancing clinical prediction models’ performances. By leveraging the rich information present in clinical text and combining it with structured EHR data, the proposed approach can improve the accuracy and robustness of predictive models. The approach has the potential to advance clinical decision support systems, enable personalized medicine, and facilitate evidence-based health care practices. Future research can further explore the application of this hybrid fusion approach in real-world clinical settings and investigate its impact on improving patient outcomes.

## INTRODUCTION

The data stored in electronic health records (EHRs) is heterogeneous and presents as varied modalities, including both structured data (e.g., vital parameters and laboratory results) and unstructured data (e.g., free-text clinical note, visit summary, and narrative information). These two modalities are both valuable for building clinically meaningful machine learning models. Clinical notes or narratives provide a detailed, personalized account of patient history and assessments, and offering a better context for clinical decision-making. Studies have demonstrated that incorporating information from clinical notes may allow for the development of better clinical prediction models, such as the prediction of mortality in critically ill patients with chronic diseases[1] and detection of adverse drug events.[2]

In natural language processing (NLP), transformer-based models have been widely applied in setting state-of-the-art benchmarks on a broad range of tasks. Bidirectional Encoder Representations from Transformers (BERT), fine-tuned to perform a specific prediction task, have led to a paradigm shift and vastly improved performance on a wide range of tasks.[3] Recent research has shown that clinical language models, pre-trained using large amounts of clinical text, outperform general-domain language models on varied clinical NLP tasks, including biomedical text mining,[4] computational phenotyping[5], and clinical question-answering.[6] These successes have been replicated in the clinical and biomedical domain via pretraining language models using large-scale clinical or biomedical corpora, then fine-tuning on a variety of clinical or biomedical downstream tasks.

While these tasks involving a single data source already show impressive performance for activity recognition, there lies a promising avenue for enhancing performance through the integration of multiple modalities.[7, 8] Ambiguities of single data sources might be resolved and the correlations between date sources could be exploited by integrating different modalities, thus improving the overall system performance. Comparing to single modality models, multimodal models can capture richer and more nuanced representations of data by incorporating multiple modalities, therefore leading to better comprehension and interpretation of complex information. By leveraging multiple modalities, multimodal models can potentially achieve higher accuracy and performance. For instance, in tasks like image captioning, combining visual and textual information can lead to more accurate and detailed captions.[9] Multimodal models enable cross-modal learning, where information from one modality can inform and improve the understanding of another modality; this can lead to better feature representations and more accurate predictions.

This study aims to develop and employ a multimodal data fusion approach to investigate whether unstructured EHR data in the form of clinical narratives contains information that could lead to better clinical prediction models when modeled together with structured EHR data.

## METHODS

### Dataset

The National Electronic Injury Surveillance System (NEISS) is a national sample of hospitals in the United States and its territories.[10] Patient information was collected from each NEISS hospital for every emergency visit involving an injury associated with consumer products. NEISS injury data are gathered from the emergency departments (ED) of approximately 100 hospitals selected as a probability sample of all 5,000+ U.S. hospitals. The system’s foundation rests on ED surveillance data, but the system also has the flexibility to gather additional data at either the surveillance or the investigation level. We choose this dataset for three reasons: 1) it contained both structured and unstructured data; 2) the length of the narratives was suitable to apply BERT-based models given the limitations of BERT models to handle long sequences; 3) it was a nationally representative dataset with low missing rates.

NEISS data contains the individual’s age, sex, race, ethnicity, injury diagnosis, affected body parts, incident locale, as well as a brief narrative description of the incident. For each case in NEISS, both structured and unstructured data were recorded. The structured data includes personal information such as age, sex, and race, and injury information such as location of injury, injured body part, fire involvement, and injury diagnosis. **Figure 1** shows several examples of the unstructured data in the narratives. A complete narrative included three parts: basic information, injury description, and diagnosis, which could be identified by terms such as ‘YOM’ (i.e., Year of Male) and ‘DX’ (i.e., diagnosis). We removed the age, and sex, and the diagnosis, and only kept the information of products and events involved in the injury.

**Figure 1.**
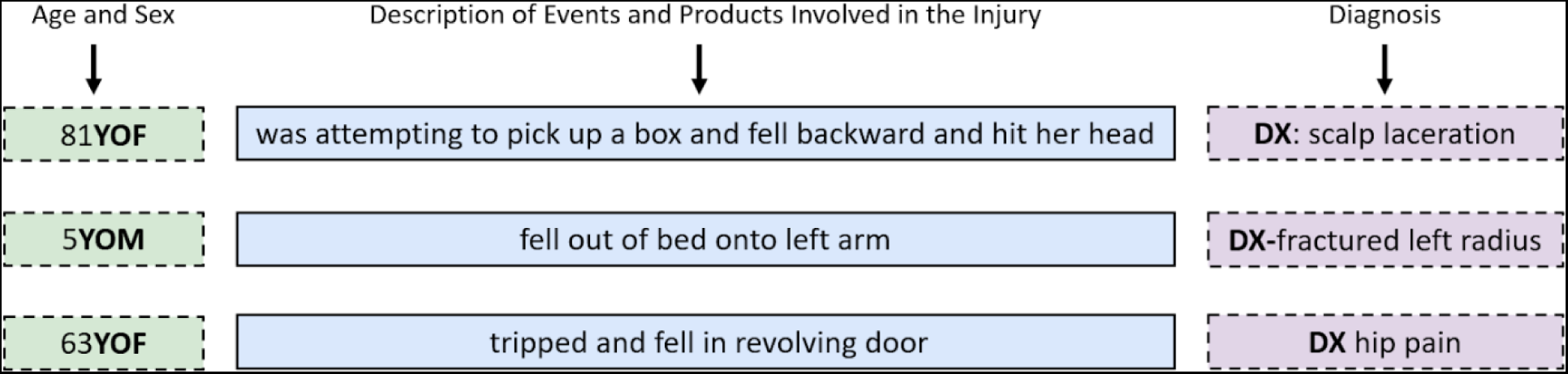
Methods pipeline and flowchart.

Figure 1 demonstrates the examples of the unstructured data in the narratives. The narratives should comply with the following format:

- Age and sex should be at the start of the narratives (i.e., 10YOM, 11MOF)
- Being descriptive and including details about the incident (i.e., who, what, why, when, where, how).
- Describing the sequence of events, alcohol/drug involvement, and the affected body part(s) in the middle of the narrative.
- Including the patient’s blood alcohol concentration/level (BAC/BAL) or breath alcohol results whenever alcohol use was associated with the incident. If a BAC/BAL or breath alcohol was not taken or recorded; the narrative should state this.
- The relevant clinician’s diagnoses should be at the end of the comment exactly as written in the ED record, and denoted with “DX:”. This abbreviation helped to distinguish the clinician’s diagnoses from other details about symptoms and complaints. If there were no clinical diagnoses in the ED record, “NO DX” were put at the end of comment.
- Quoting verbatim the words actually used within the ED record.

Examples of comments following these guidelines:

- 60YOF woke up on floor, unsure if fell from bed or if woke up and fell, heavy alcohol use night before. +5cm laceration thru all subcutaneous layers. BAC 0.19%. DX: laceration of forehead.
- 25YOM cut his hand when picking up broken glass from a bowl that dropped on the floor. DX: laceration of superficial palmar arch of right hand; laceration of left index finger without foreign body.
- 80YOF was vacuuming when walking from bedroom to hallway and caught her foot on an extension cord, falling to floor and heard pop in hip. Admitted. DX: fall; left femoral neck fracture.

In this study, the prediction outcome was the injury diagnosis. The structured personal information, structured injury information, and unstructured narrative information were used as model inputs. Records with missing values of diagnosis and/or without ‘DX’ in the narratives were dropped from the raw dataset.

### Language model

The complex nature of unstructured narrative data, primarily characterized by long sentences with intricate word relationships, calls for a potent language model capable of decoding semantic information to extract diagnosis-related data. Recent advancements have witnessed the application of pre-trained transformer-based language models, such as BERT [3] and RoBERTa (A Robustly Optimized BERT Pretraining Approach),[11] in text analysis. RoBERTa took the BERT architecture a step further by experimenting with different training approaches and hyperparameters, leading to improved performance. These models, noted for their commendable ability to capture contextual information in long sequence data through attention mechanisms, are becoming more prevalent. In the realm of health care and clinical fields, language models enriched with clinical knowledge like Clinical-BERT [12] and Clinical-Longformer[13] have emerged. ClinicalBERT is a modified BERT model to clinical corpora to address the challenges of clinical text and the representations are learned using medical notes and further processed for downstream clinical tasks.[12] Clinical-Longformer is a clinical knowledge enriched version of Longformer that was further pre-trained using clinical notes; it can allow up to 4,096 tokens as the model input. [13] These models, pre-trained on clinical texts as well as resources like BookCorpus, are engineered to outperform their counterparts in handling clinical downstream tasks.

### Multimodal modality fusion

Unstructured data and structured data can be considered as two data modalities. Unstructured data, such as the descriptions of injury causes and symptoms in the clinical narratives, typically contains abundant information. However, the integration of varying content types within a single sentence can present certain challenges for semantic understanding and information extraction. Contrarily, structured data (e.g., affected body parts) offers a more organized format, where diverse information types such as incident locations or injured body parts are clearly categorized under distinct classifications. Unimodal models leverage either structured or unstructured data for inference and prediction, whereas multimodal models strive to integrate both types to enhance predictive performance.

In multimodal analysis, there are two common modality fusion strategies: early fusion and late fusion.[14, 15] Early fusion, a data-level fusion method, amalgamates data from diverse modalities prior to model input. While it is noted for its superior ability to capture the interaction of different modalities, it necessitates a multimodal encoder capable of processing multimodal inputs, making the application of pre-trained unimodal models challenging in this context.[16] Different from early fusion, late fusion is an embedding level fusion approach, which first uses independent encoders to process different modalities, and then fuses embeddings from multiple encoders in a post-processing model.[16, 17] Pre-trained unimodal models can be easily applied as independent encoders in a late fusion. With its flexibility of leveraging pre-trained unimodal models, late fusion is more commonly used in multimodal analysis field.[18]

To use both unstructured narrative text and structured patient information to predict injury diagnosis, we converted structured data into unstructured text for effective early fusion. While leveraging powerful pre-trained language models for processing text data, a late fusion model was first proposed as the baseline model. As shown in Figure 2 **(i)**, this model utilized a pre-trained language model as the text encoder and a randomly initialized fully-connected block as the structured data encoder. The embeddings from these two encoders are then concatenated and subsequently fed into the post-prediction layers to predict the final diagnosis. However, this late fusion model has several disadvantages: 1) the structured data encoder, not having been pre-trained, might exhibit instability and lack of generalization during the training phase; and 2) the interaction between structured data and unstructured text information could be constrained by the use of independent encoders, thus potentially undermining the overall model performance.

**Figure 2.**
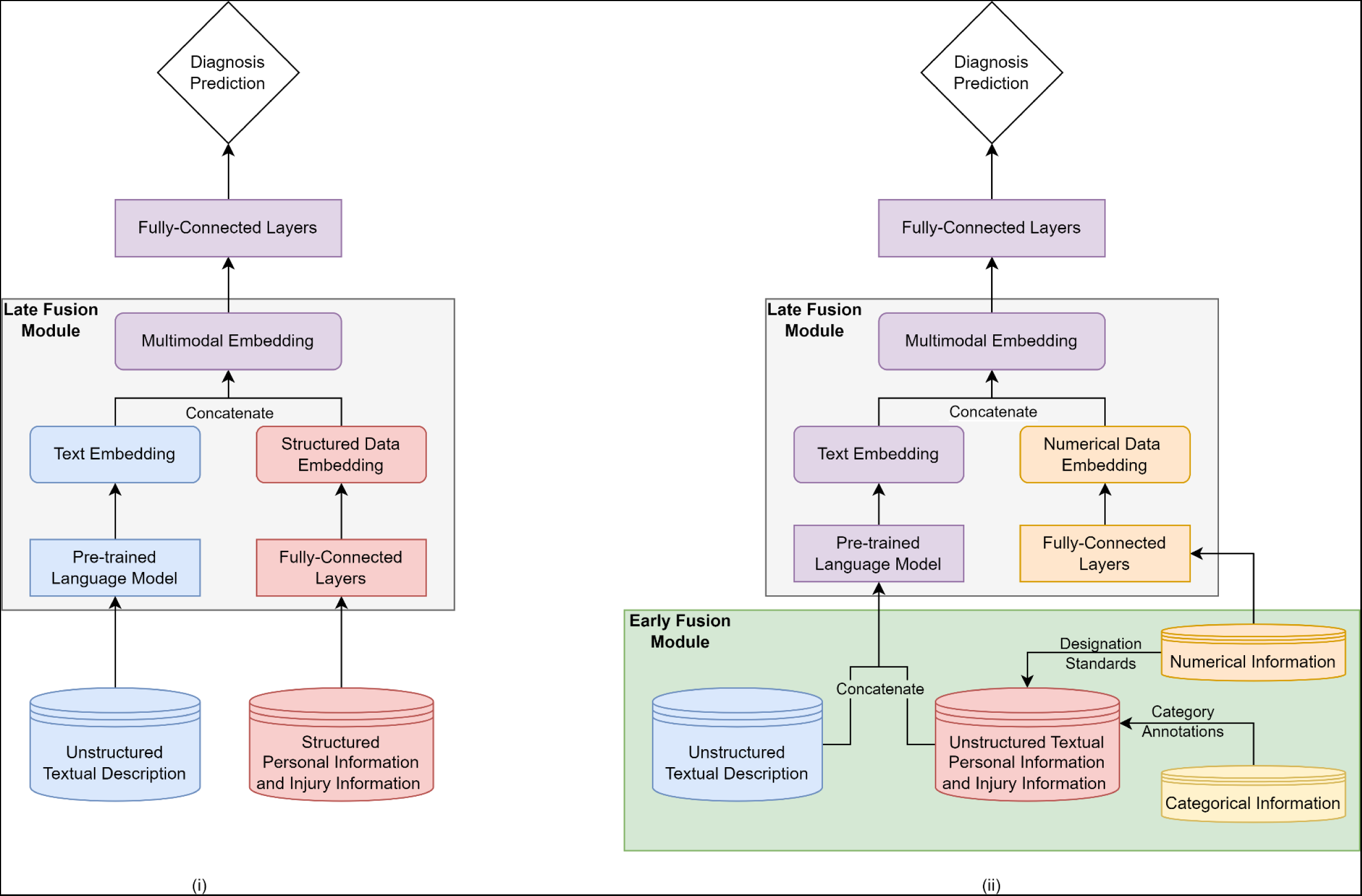
The frameworks of late fusion baseline(i) and proposed hybrid fusion method(ii).

In an attempt to leverage the benefits of both early and late fusion strategies while surmounting their limitations, we proposed a hybrid fusion approach for multimodal injury diagnosis. As illustrated in Figure 2 **(ii)**, this system amalgamated the techniques of early and late fusion. Given that structured data bear significant semantic information and possess the potential to be interpreted by the language model, we integrated an early fusion module to incorporate structured data into raw narrative contents. Structured data were initially transformed into unstructured words based on their semantic meanings and processed jointly with unstructured text by the pre-trained language model. To elaborate, categorical variables were converted into semantic annotations; for instance, a “sex” value of 1 was translated to its original definition “male”. Numerical variables were transformed into cross-grained categories in accordance with commonly applied designation standards. For instance, ages ranged from 0-1 was defined as infant, 1-12 were children, 12-19 were teenager, 19-45 were young adults, 45-65 was middle age adults, and over 65 were old adults. However, this method might fall short in capturing fine-grained regression information. To exploit the fine-grained numerical information in the structured data, numerical variables were directed into a fully-connected encoder and subsequently fused with the output of the text encoder via a late fusion strategy. Unstructured words from narratives were concatenated with unstructured data and then fed into a pre-trained language model as text encoder. The resultant concatenated embedding was then fed into a post-processing block for the final diagnosis prediction. This strategy utilized pre-trained language models to understand semantic information in the structured data and added more interaction within structured data, and between structured data and unstructured data by self-attention mechanism in the transformer-based language model. In addition, inspired by a location-aware sound event detection model,[19] we used the language model to encode categorical location variable to handle unseen sound scenes in test data, which could also potentially handle unseen categories in the structured data.

### Experimental Setup

To mitigate unevenly distributed injury categories in this dataset, a random sampling approach was first applied in each diagnosis category for each year. For categories with more than 1,000 records, only 1,000 records were randomly sampled; for categories with less than 1,000 records, all records were sampled. To evaluate and compare model performance, a stratified random sampling based on categories distribution was applied to split training, validation, and testing dataset with an 80:10:10 ratio.

To compare the effects of unstructured and structured data for injury prediction and validate the effectiveness of multimodal fusion, we compared the unimodal systems using either unstructured or structured data with the multimodal systems using both modalities. For multimodal systems, the late fusion strategy was set as the baseline and compared with the proposed hybrid fusion method.

For the language module used to extract information from the unstructured data, the pre-trained BERT model was employed in both the unimodal system with unstructured data and the multimodal systems. For the proposed hybrid fusion approach, we expanded our comparison set by integrating three additional language models alongside BERT: RoBERTa, Clinical-BERT, and Clinical-LongFormer.

To further improve the prediction performance on unbalanced categories with less data, we minimized focal loss function (as shown in function below) [20], which was designed to address class imbalance issues.

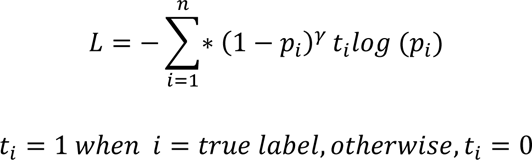

Where *n* denotes the number of categories, *p_i_* denotes the probability distribution of the prediction, the *t_i_* denotes the real probability distribution, and the 𝛾 denotes the focusing parameter.

For evaluation, we used the Top 1 injury prediction accuracy on the validation dataset to identify the best-performed model checkpoint. We assessed and compared the best testing performance using multiclass classification metrics, including Top 1 injury’s accuracy, macro F1-score, and weighted macro F1-score. Because there were 24 injury diagnosis categories in this multiple classification task, which made it challenging to achieve an extremely high accuracy for the Top 1 injury. In addition, the causes and characteristics of some injuries might be similar or intercorrelated, which made it also difficult to distinguish between these categories. Therefore, we introduced Top 2 and Top 3 injuries’ accuracy metrics into our evaluation framework, reflecting real-world application scenarios.

## RESULTS

**Table 1** presents the sample characteristics for the overall study population. There were 1,725,802 patients in this study, and 777,143 (45.0%) were females. The mean (SD) age was 30.1 (26.6) years old. The majority patients were White (52.5), and 25% of the patients were not Hispanic.

**Table 1.** Sample characteristics for the overall study population and by sex.

| Variable | Total (N=1,725,802) | Male (N=948,659) | Female (N=777,143) | P-value |
| --- | --- | --- | --- | --- |
| Age, mean (SD) | 30.1 (26.6) | 26.7 (24.2) | 34.2 (28.8) | <0.001 |
| Race, N (%) |  |  |  | <0.001 |
| White | 905,423 (52.5) | 485,828 (51.2) | 419,595 (54.0) |  |
| Black | 259,704 (15.1) | 148,917 (15.7) | 110,787 (14.3) |  |
| Asian | 29,682 (1.72) | 16,536 (1.74) | 13,146 (1.69) |  |
| American Indian/Alaska Native | 5,674 (0.3) | 3,089 (0.3) | 2,585 (0.3) |  |
| Native Hawaiian/Pacific Islander | 2,194 (0.1) | 1,300 (0.1) | 894 (0.1) |  |
| Other* | 103,597 (6.0) | 60,750 (6.4) | 42,847 (5.5) |  |
| Not stated | 419,528 (24.3) | 232,239 (24.5) | 187,289 (24.1) |  |
| Hispanic, N (%) |  |  |  | <0.001 |
| Yes | 130,411 (7.6) | 76,253 (8.0) | 54,158 (7.0) |  |
| No | 430,735 (25.0) | 229,581 (24.2) | 201,154 (25.9) |  |
| Not stated | 1,009,433 (58.5) | 557,846 (58.8) | 451,587 (58.1) |  |
| Missing | 155,223 (9.0) | 84,979 (9.0) | 70,244 (9.0) |  |

\*Other: more than one race (e.g., multiracial, biracial).

Figure 3 demonstrates the distribution of body part categories. Subsets of burns such as scald, radiation, and chemical burns were merged into one category. The highest frequent body part categories include head (19.29%), face (10.09%), finger (9.15%), lower trunk (6.25%), and ankle (5.67%).

**Figure 3.**
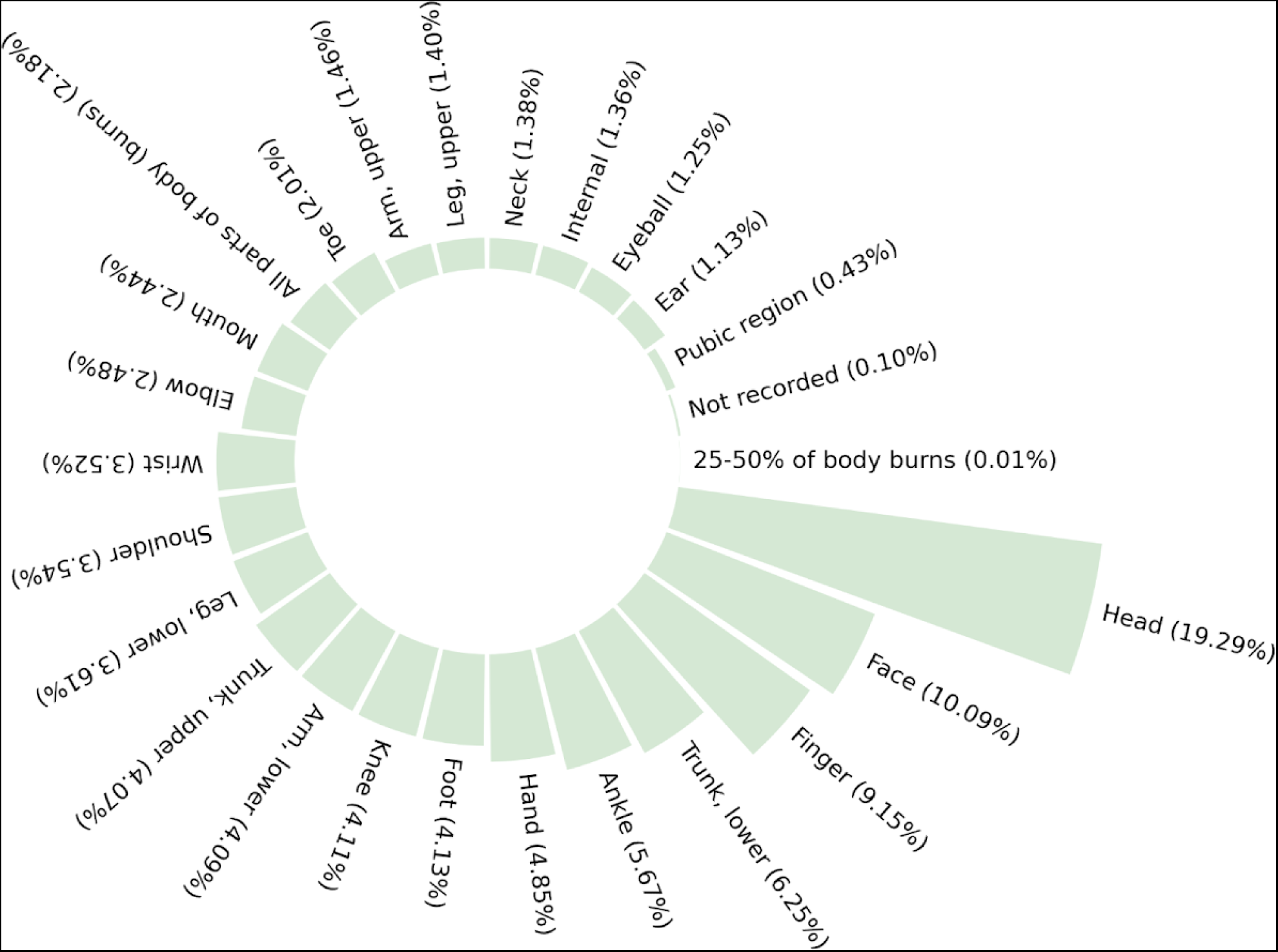
The distribution of body part categories.

Figure 4 demonstrates the distribution of all the diagnosis categories. The highest frequent injury diagnosis include laceration (20.38%), fracture (19.38%), contusions/ abrasions (15.15%), strain or sprain (14.47%), and internal organ injury (12.22%).

**Figure 4.**
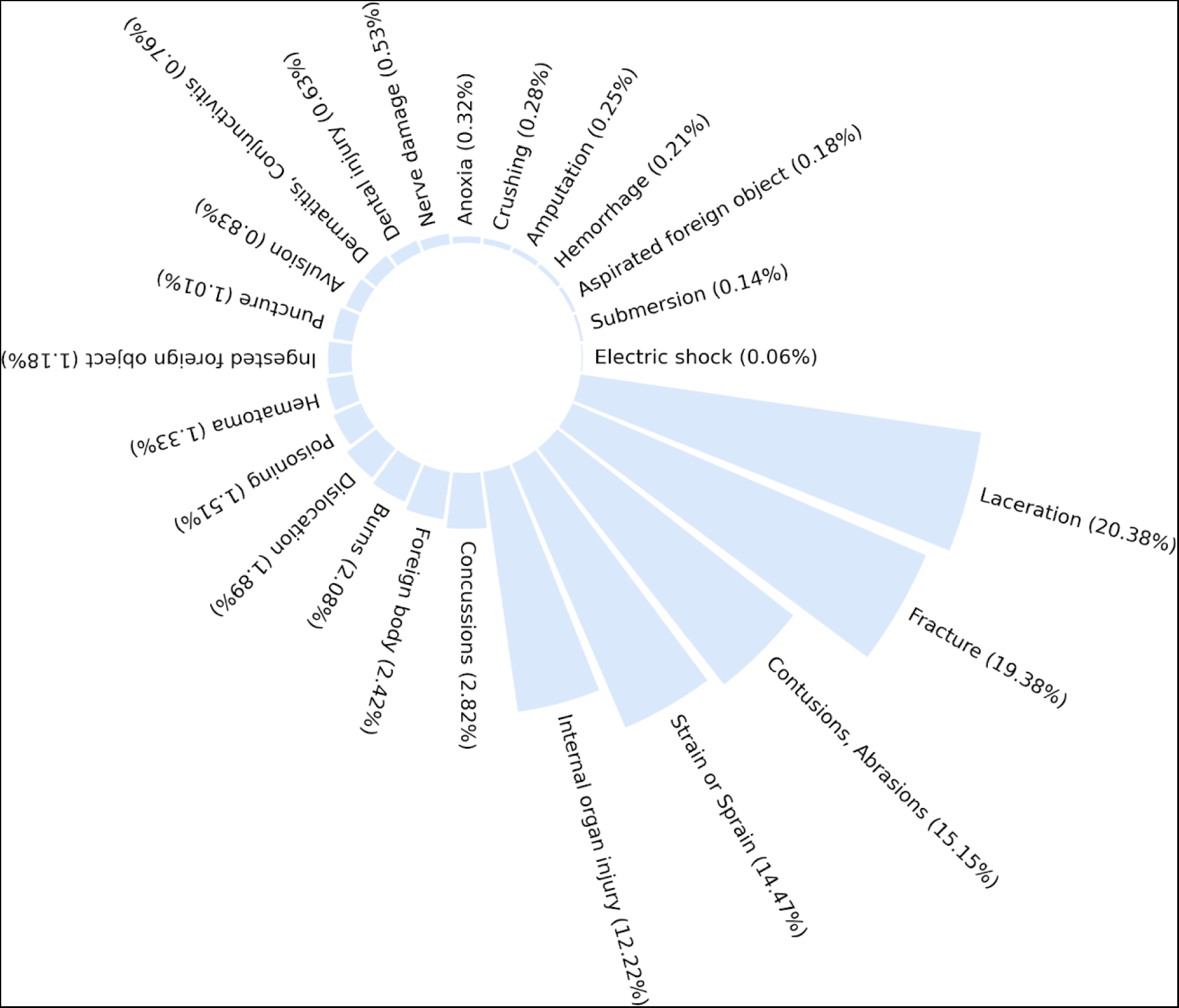
The distribution of diagnosis categories. Subsets of burns such as scald, radiation, and chemical burns were merged into one category.

**Table 2** shows the performance of the proposed models. RoBERTa model with hybrid fusion has the highest accuracy and weighted Macro F1 Score for predicting the Top 1 injury as well as the accuracy to predict the Top 2 and Top 3 injuries.

**Table 2.** The performance of models.

| Method | Language Module | Accuracy for Top 1 injury | Macro F1 Score | Macro F1 Score (Weighted) | Accuracy for Top 2 injury | Accuracy for Top 3 injury |
| --- | --- | --- | --- | --- | --- | --- |
| Unimodal (Structured) | None | 50.08 % | 42.02 % | 53.85 % | 67.04 % | 76.33 % |
| Unimodal (Unstructured) | BERT | 67.48 % | 69.42 % | 67.85 % | 81.31 % | 87.48 % |
| Late Fusion | BERT | 70.77 % | 73.02 % | 70.86 % | 84.01 % | 89.71 % |
| Proposed Hybrid Fusion | BERT | 74.70 % | 76.33 % | 75.07 % | 88.13 % | 93.43 % |
| Proposed Hybrid Fusion | RoBERTa | **75.00 %** | 76.61 % | **75.44 %** | **88.50 %** | **93.54 %** |
| Proposed Hybrid Fusion | Clinical-BERT | 74.85 % | 76.55 % | 75.33 % | 88.12 % | 93.40 % |
| Proposed Hybrid Fusion | Clinical-LongFormer | 74.88 % | **76.81 %** | 75.18 % | 88.03 % | 93.32 % |

## DISCUSSION

The primary aim for this paper was to study how to best incorporate information from clinical notes and structured patient information in a multimodal prediction system. We evaluated both unimodal and multimodal systems for the injury diagnosis prediction task. The best-performing model was RoBERTa, with an accuracy of 75.00% for predicting the Top 1 injury. The experiment results indicated that the multimodal systems outperformed unimodal systems, and the proposed hybrid fusion strategy further improved the performance of multimodal fusion model. We evaluated the data fusion strategies for leveraging a clinical BERT model in a multimodal setup, which allowed for creating richer text representations that facilitate learning cross-modal interactions.

The motivation for employing a multimodal modeling approach was to investigate whether unstructured data in the form of clinical narratives contained information that could lead to better clinical prediction models when modeled together with structured data. The experimental results showed that the multimodal model substantially outperformed the unimodal model based on the sole use of structured data or unstructured data on this task. In fact, the unimodal model based on only clinical narratives outperformed a unimodal model based on only structured data. This result demonstrates that clinical notes can introduce valuable information for predicting the diagnosis of injury. The results also showed that the proposed multimodal model, based on multimodal fine-tuning via the hybrid fusion strategy, clearly outperformed the late fusion system which modeled the different modalities separately and then combined their global embeddings for the final prediction. This can be seen as an indication that the proposed multimodal model is able to capture cross-modal interactions between the structured and unstructured data.

Given the limitations of BERT for clinical applications, such as incompletely optimized training strategy, non-clinical training data and long data processing capabilities. We conducted a comparison study of the hybrid fusion system with different pre-trained language encoders. The results in **Table 2** demonstrate that the performance of BERT can be further improved by utilizing BERT-based model. RoBERTa, like BERT, is a type of unsupervised, transformer-based model. RoBERTa was pre-trained on a massive corpus of text data in a masked language modeling task, and the resulting model could be fine-tuned on specific NLP tasks like classification, entity recognition, and more. Interestingly, the system with RoBERTa that was pre-trained on non-clinical data achieved better performance than systems with the language model pre-trained on additional clinical data. The reasons leading to this phenomenon may include: 1) RoBERTa used larger batch sizes during pretraining, which helped it learn better representations; 2) RoBERTa was trained for more epochs compared to BERT, allowing it to capture more complex language patterns; 3) while BERT used static masking of words during pretraining, RoBERTa employed dynamic masking, where a new set of masked words was sampled for each epoch of training; 4) RoBERTa was trained on a larger corpus of text data, incorporating data from books, articles, and more sources, which enhanced its ability to understand a wider range of language; and 5) RoBERTa systematically tuned hyperparameters like learning rate, batch size, and training duration to achieve optimal performance. In addition, RoBERTa performed better compared to Clinical-BERT and Clinical-LongFormer using the proposed hybrid fusion for predicting the injury diagnosis in this study. The reasons may include: 1) the dataset used in this study was large, eliminating the need to rely on clinical data for pre-training, and 2) the clinical data used for pre-training differed from the textual data in this study (the textual data in MIMIC III often lacks detailed symptom descriptions and causes of injury). Consequently, the knowledge acquired from this type of clinical data offered minimal assistance in the prediction of injuries in our study.

In terms of the application scenarios, we utilized the nationally representative injury data across eleven years to maximize the generalizability. Injuries are the leading cause of death and disability in the United States. In 2021, 224,935 patients died as a result of unintentional injury, as the fourth course of death.[21] Characterizing the severity and mechanism of these injuries is important for prognostication, management, and participation in quality improvement and clinical trials.[22] Automatic methods are needed to reduce human labor related to manual chart review to improve throughput and enable comprehensive risk scoring on all injury admissions.[23] Injury prediction can improve allocation of resources, stratification of patients by risk and inform decision making. Results assist with prediction about the patients’ current state, but system properties limit forecasting far in the future with any degree of accuracy. For the future work, we will build an integrated system that collects the clinical information as well as patient-generated health data, including the brief narrative description of the cases. This system will be also built in the clinical decision support system to 1) produce pre-diagnosis, 2) triage patients, and 3) provide better decisions for clinicians. By identifying patients who are at higher risk of certain injuries, clinicians can prioritize interventions and allocate resources such as staff time, equipment, and facilities to provide targeted care and monitoring. This system can also identify individuals at higher risk of specific injuries based on various risk factors from the structured data as well as unstructured data, such social determinates of health. Clinicians can use this information to educate patients about potential risks and provide personalized recommendations to prevent injuries; this could include advice on physical activity modifications, safety precautions, and lifestyle changes.[24] Furthermore, this system can be integrated into telehealth platforms,[25] allowing clinicians to remotely monitor patients and provide timely interventions, which is particularly important for patients with chronic conditions or those at high risk of specific injuries and need immediate treatment.[26]

### Limitation

This study has a few limitations. First, the success of the proposed hybrid fusion approach heavily relied on the availability and quality of multimodal clinical data. The number of features was small in the NEISS datasets. Second, even though NEISS datasets are nationally cross-sectional, the performance of the proposed approach may vary across different clinical settings or health care institutions. In the future, we will validate this approach in other datasets with diverse data modalities. Third, integrating multiple modalities and applying natural language processing techniques may increase the computational complexity and resource requirements of the proposed approach; this may limit its scalability, particularly when dealing with large-scale datasets or real-time prediction scenarios. The proposed approach may need high computational resources and the time-consuming processing may restrict the practical implementation of the hybrid fusion approach in resource-constrained healthcare environments.

## CONCLUSION

This study developed a multimodal model that combined both structured and unstructured data. We also studied how to make best use of pre-trained language models in a multimodal setup for hybrid data fusion. We applied the multimodal model for predicting injury upon hospital admission using a nationally representative dataset and evaluated unimodal and multimodal prediction systems. We also explored various pre-trained language models in the proposed hybrid fusion system and the experiment results revealed the advantages of RoBERTa in handling non-standard clinical notes. The experimental results indicated that clinical prediction models can be improved by including multimodal data modality and utilizing the data hybrid fusion framework with the pre-trained language mode for effective multimodal data fusion. The proposed system achieved promising accuracy in injury prediction and could be potentially used in applications such as injury predictions in outpatient clinics or at the emergency department.

## FUNDING

None.

## AUTHOR CONTRIBUTIONS

J.Y. conceived of the idea, designed the study, contributed to the analyses, and drafted the manuscript. J.H. contributed to the analyses and drafted the manuscript. All the authors contributed to the interpretation of the results and revision of the manuscript. All the authors read and approved the final version of the manuscript.

## DATA AVAILABILITY STATEMENT

All data referred to in the manuscript are publicly available. The relevant code and analyses are available at: https://github.com/haidog-yaqub/Clinical_HybridFusion.

## CONFLICT OF INTEREST STATEMENT

None.

## Notes

### Competing Interest Statement

The authors have declared no competing interest.

### Funding Statement

This study did not receive any funding

